# Evaluating the Diagnostic Accuracy of Anti- Zinc Transporter 8 Antibodies in Type 1 Diabetes: A Systematic Review and Meta-Analysis

**DOI:** 10.1101/2024.06.07.24308627

**Authors:** Luís Jesuíno de Oliveira Andrade, Gabriela Correia Matos de Oliveira, Roseanne Montargil Rocha, Alcina Maria Vinhaes Bittencourt, Túlio Matos David, Luís Matos de Oliveira

## Abstract

**Background:** The association between autoantibodies and the risk of type 1 diabetes mellitus (T1DM) is well established. However, there is still a lack of quantitative insight into the role of anti-zinc transporter 8 (anti-ZnT8) antibodies and their efficacy in T1DM diagnosis.

**Objective:** To conduct a systematic review and meta-analysis on the association between anti-ZnT8 autoantibodies and the risk of T1DM.

**Methods:** Relevant studies were retrieved from the PubMed database and analyzed using a fixed or random-effects model.

**Results:** Out of 211 screened articles, 23 studies were selected and a total of 14,172 patients were included in the analysis. Our pooled analysis revealed that anti-ZnT8 autoantibody expression was significantly associated with an increased risk of T1DM development in both children and adults [OR 1.14 (95% CI: 1.12-1.16); P = 0.07].

**Conclusion:** Our systematic review and meta-analysis provides robust evidence supporting a significant association between anti-ZnT8 autoantibody positivity and an increased risk of T1DM development.

## INTRODUCTION

Zinc transporter 8 (ZnT8) antibodies have emerged as a promising biomarker for the diagnosis of type 1 diabetes mellitus (T1DM), a chronic autoimmune disease characterized by the destruction of insulin-producing beta cells in the pancreas, leading to insulin deficiency and hyperglycemia.^1^ ZnT8 is a transmembrane protein predominantly expressed in pancreatic beta cells and plays a crucial role in regulating intracellular zinc homeostasis.^2^ The presence of ZnT8 autoantibodies, specifically directed against intracellular ZnT8, is considered a hallmark of T1DM, often detectable years before clinical manifestation.^3^

T1DM remains a global health challenge, affecting millions of individuals worldwide, with a peak incidence in childhood and adolescence.^4^ The disease is characterized by the progressive loss of beta cells, resulting in insulin deficiency and dysregulated blood glucose levels.^5^ While insulin therapy serves as the mainstay of T1DM management, early diagnosis is crucial for timely intervention and potential disease prevention.^6^ The identification of individuals at risk for T1DM holds immense promise for early intervention strategies, aiming to halt or delay beta cell destruction and preserve insulin secretion.

The association between ZnT8 antibodies and T1DM has been extensively studied, demonstrating a strong correlation between their presence and the development of the disease.^7^ ZnT8 autoantibodies are detectable in the serum of individuals with T1DM, often preceding clinical symptoms by several years, making them valuable tools for early diagnosis and risk stratification.^8^ The presence of these antibodies has been shown to have high specificity for T1DM, with minimal detection in other autoimmune conditions.^9^ Additionally, ZnT8 autoantibody levels have been shown to correlate with the progression of beta cell destruction, providing a potential biomarker for monitoring disease activity.^10^

Despite the promising role of ZnT8 antibodies in T1DM diagnosis, there is still a need for a comprehensive assessment of their diagnostic accuracy.^11^ The existing literature on ZnT8 antibodies is characterized by a substantial heterogeneity in study design, methodology, and patient populations, making it challenging to draw definitive conclusions about their overall performance.^12^ A systematic review and meta-analysis are warranted to synthesize the available evidence and provide a robust evaluation of the diagnostic accuracy of ZnT8 antibodies for T1DM.

Therefore, the objective of this manuscript is to conduct a systematic review and meta-analysis of studies that have investigated the diagnostic accuracy of ZnT8 antibodies for T1DM. We will systematically identify, appraise, and synthesize relevant studies to estimate the pooled sensitivity, specificity, and positive and negative predictive values of ZnT8 antibodies for T1DM diagnosis. Additionally, we will explore potential sources of heterogeneity between studies and investigate factors that may influence the performance of ZnT8 antibodies. The findings of this review will provide valuable insights for clinicians and researchers seeking to utilize ZnT8 antibodies in the diagnosis and management of T1DM.

## METHODOLOGY

### Study identification and selection

A comprehensive search of the PubMed database will be conducted to identify relevant studies for this systematic review and meta-analysis. The search strategy will be developed using a combination of keywords and MeSH (Medical Subject Headings) terms related to ZnT8 antibodies, T1DM, diagnosis. The following search terms will be used: (znt8[All Fields] AND (“antibodies”[MeSH Terms] OR “antibodies”[All Fields])) AND (“diabetes mellitus, type 1”[MeSH Terms] OR “type 1 diabetes mellitus”[All Fields] OR “type 1 diabetes”[All Fields]).

The research strategy involved key electronic databases including PubMed from their 2007 Jan - 2024 June^13^.

The search will be restricted to studies published in English, and no date limits will be applied. Additional studies will be identified by screening the reference lists of retrieved articles and relevant reviews.

### Eligibility criteria

Studies will be included in the review if they meet the following eligibility criteria: The study is a primary or secondary study that investigates the diagnostic accuracy of ZnT8 antibodies for T1DM. The study reports data on sensitivity, specificity, positive and negative predictive values, or other relevant diagnostic measures for ZnT8 antibodies. The study is available in full text.

Studies will be excluded if they meet any of the following criteria: The study is a case report, review, editorial, letter to the editor, or other non-research article. The study does not report data on the diagnostic accuracy of ZnT8 antibodies. The study is not available in full text.

### Data extraction

Two reviewers will independently extract data from the included studies using a standardized data extraction form. The data extraction form will include information on study characteristics, patient characteristics, study design, methodology, and diagnostic outcomes. Any discrepancies in data extraction will be resolved through discussion or by consulting a third reviewer.

### Quality assessment

The quality of the included studies will be assessed using the Quality Assessment of Diagnostic Accuracy Studies 2 (QUADAS-2) tool.^14^ The QUADAS-2 tool is a checklist of 14 items that assesses the risk of bias in studies of diagnostic accuracy. Each item will be scored as “low risk,” “high risk,” or “unclear risk.” The overall quality of each study will be judged as “good,” “fair,” or “poor” based on the number of high-risk and unclear risk items.

### Data synthesis

Pooled estimates of sensitivity, specificity, positive and negative predictive values, and diagnostic odds ratios (OR) will be calculated for ZnT8 antibodies using a random-effects meta-analysis model. The I^2^ statistic will be used to assess heterogeneity between studies. If significant heterogeneity is present, subgroup analyses will be conducted to explore potential sources of heterogeneity.

Assessment of the quality of evidence using the metaHUN: a web tool http://softmed.hacettepe.edu.tr/metaHUN). (^15^

### Sensitivity analysis

A sensitivity analysis will be conducted to assess the robustness of the meta-analysis findings by excluding studies with high or unclear risk of bias.

### Publication bias

Publication bias will be assessed using funnel plots and Egger’s test.

### Additional analyses

Additional analyses will be conducted to investigate the impact of study characteristics, such as patient age, disease duration, and assay type, on the diagnostic accuracy of ZnT8 antibodies.

## CONSIDERATIONS REGARDING RESEARCH ETHICS

Since this investigation constitutes a secondary analysis of existing data, formal ethical approval was deemed unnecessary. This stems from the fact that secondary analyses, by their very nature, do not entail direct engagement with human participants. Consequently, they introduce no new ethical concerns beyond those inherent to the original data collection process.

## RESULTS

To ensure transparency and methodological rigor in our study selection, we implemented a comprehensive and structured literature search strategy aligned with the PRISMA^16^ framework (Figure 1). This process commenced with the identification of 211 potentially relevant articles using a meticulous search strategy that incorporated well-defined keywords and Boolean operators within relevant databases. Following an initial screening based on titles and abstracts, 82 articles were deemed worthy of a more in-depth evaluation. This subsequent phase involved a thorough examination of both abstracts and full-text manuscripts to meticulously assess their alignment with our research focus: the safety and therapeutic potential of topical insulin treatments in ophthalmology. Ultimately, the rigorous application of our predefined eligibility criteria yielded a final selection of 23 pivotal studies for data extraction and synthesis.^17-39^

**Figure 1.**
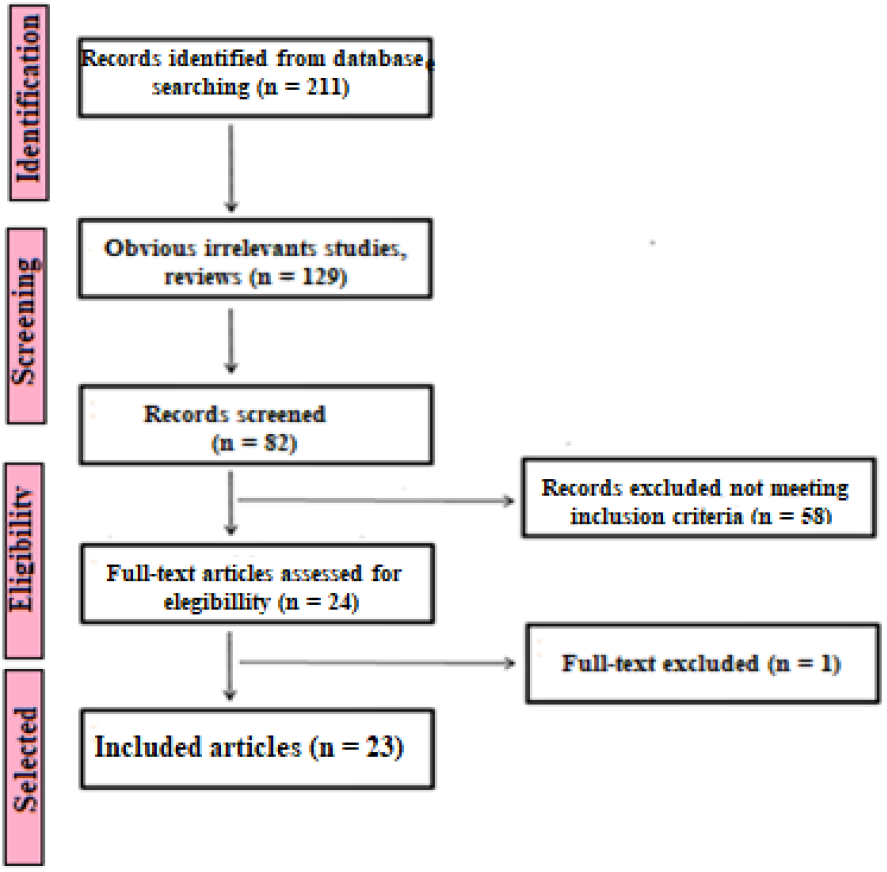
Flowchart of the selection process for the 23 studies included.

This systematic literature review aimed to comprehensively evaluate investigated the diagnostic accuracy of ZnT8 antibodies for T1DM. The twenty-two studies ultimately selected encompassed a diverse range of methodological approaches. These included case-control (n=2), retrospective (n=3), and cross-sectional (n=18) designs. The research originated from a variety of geographical locations and included studies with sample sizes ranging from 16 to 3,165 participants. After general evaluation of the studies, a combined total of 23 studies fulfilled our inclusion criteria for quantitative analysis within the systematic review.

### Primary features of the chosen research works

#### Source 1

Basu M, et al.^17^; Objective: Assessing the autoantibody profile in children with T1DM; Type of study: cross-sectional study; Participants: 92 subjects with T1DM (33 males, 59 females); Results: ZnT8 antibody were present in 20.65% subjects; Conclusion: “T1DM is associated with a high prevalence of autoantibodies and antibody negative T1DM is rare”.

#### Source 2

Lounici Boudiaf A, et al.^18^; Objective: Evaluate the prevalence of islets cells antibodies, glutamic acid decarboxylase, islet antigen type 2, insulin autoantibodies, and ZnT8 antibodies in young Algerian patients with T1DM; Type of study: cross-sectional study; Participants: 160 patients (74 males and 86 females) between 1 and 35 years old, diagnosed with type 1 diabetes; Results: ZnT8ab was positive in 70.3% females and 10.7% in males; Conclusion: “ZnT8ab is a good tool for differential diagnosis of T1DM”.

#### Source 3

Reference: Zecevic-Pasic L, et al.^19^; Objective: to analyze presence of T1DM-related autoantibodies; Type of study: cross-sectional study; Participants: 67 patients with T1DN (40 male and 27 female); Results: 36 (53,7%) cases were positive for Zn-T8; Conclusion: “Zn-T8 antibodies is the most frequently detected and is an important serological marker of T1DM”

#### Source 4

Reference: Elmaoğullari S, et al.^20^; Objective: to investigate the prevalence of ZnT8A in Turkish children with new onset T1DM; Type of study: cross-sectional study; Participants: 84 patients diagnosed with T1DM and 50 healthy children without any autoimmune diseases; Results: ZnT8A positivity was detected in 58% of the patients with new onset T1DM and 8% of the control group; Conclusion: “ZnT8 measurement should be more widespread for clarifying the etiology in T1DM”.

#### Source 5

Reference: Bhola S, et al.^21^; Objective: To establish the frequency of ZnT8 autoantibodies in black South Africans diagnosed with T1DM, and investigate potential correlations between ZnT8 autoantibody positivity, age at diagnosis, and duration of the disease; Type of study: cross-sectional study; Participants: Patients with T1DM (n = 183) and controls (n = 49) Results: The prevalence of ZnT8 autoantibody positivity was 17.5 % (32 of 183) in participants with T1DM and 27.3 % (6 of 22) in newly diagnosed participants; Conclusion: “The greater the numbers of autoantibodies present in an individual the earlier the age at diagnosis”.

#### Source 6

Reference: Thewjitcharoen Y, et al.^22^; Objective: To describe the characteristics of long-standing T1DM in Thai patients and assess residual beta-cell function with status of pancreatic autoantibodies; Type of study: cross-sectional study; Participants: 20 patients (males 65%); Results: The prevalence rate of anti-ZnT8 was 10%; Conclusion: “Endogenous insulin secretion persists in some patients with long-standing T1DM”.

#### Source 7

Reference: Andersson C, et al.^23^; Objective: To determine the diagnostic accuracy and associations among autoantibodies targeting the three different variants of ZnT8A at position 325, human leukocyte antigen-DQ, and autoantibodies against glutamic acid decarboxylase, insulinoma-associated protein 2, and insulin; Type of study: cross-sectional study; Participants: 3,165 patients with T1DM; Results: ZnT8A was found in 65% of the patients; Conclusion: “Analysis of ZnT8A increased the diagnostic sensitivity of islet autoantibodies for T1DM”.

#### Source 8

Reference: Andersson C, et al.^24^; Objective: To evaluate the diagnostic sensitivity enhancement for T1DM through the combination testing of ZnT8RWQ autoantibodies, GAD65 autoantibodies, insulinoma-associated protein 2 autoantibodies, insulin autoantibodies, and islet cell cytoplasmic autoantibodies with human leukocyte antigen; Type of study: cross-sectional study; Participants: 3165 patients with T1DM; Results: ZnT8RWQA autoantibodies was found in 65% (449/686) of the patients; Conclusion: “The results suggest that ZnT8RWQA is a necessary complement to the classification and prediction of T1DM”.

#### Source 9

Reference: Su YT, et al.^25^; Objective: to report the prevalence, diagnostic utility, and clinical characteristics of ZnT8A in children with T1DM; Type of study: cross-sectional study; Participants: 268 children (130 boys, 138 girls) newly diagnosed with T1DM; Results: ZnT8A was detected in 117 patients (43.7 %); Conclusion: “ZnT8A testing can diagnose up to 12 % more patients with T1DM along with three other antibodies”.

#### Source 10

Reference: Fakhfakh R, et al.^26^; Objective: to evaluate the relationships between ZnT8-Ab, ZnT8 coding gene (SLC30A8) promoter polymorphism, and T1DM risk in newly diagnosed children; Type of study: cross-sectional study; Participants: ZnT8-Ab were measured in the serum of T1DM newly affected children (n = 156); Results: ZnT8-Ab was detected in 66/156 (42.3%); Conclusion: “ZnT8-Ab appears as a relevant diagnostic marker for T1DM in Tunisian children”.

#### Source 11

Reference: Fuentes-Cantero S, et al.^27^; Objective: to evaluate the diagnostic efficiency of ZnT8 autoantibodies in diagnosis of T1DM in pediatric patients; Type of study: retrospective study; Participants: 80 patients under 16 years of age with suspected T1DM; Results: ZnT8A obtained the most significantly global diagnostic accuracy (0.75); Conclusion: “The results obtained indicate a higher efficiency of anti-ZnT8 autoantibodies for the diagnosis of T1DM in pediatric patients”.

#### Source 12

Reference: Baumann K, et al.^28^; Objective: to evaluate autoantibodies anti-ZnT8 in schoolchildren from the general population and in people with autoimmune diabetes; Type of study: cross-sectional study; Participants: 137 schoolchildren with T1DM, 102 people at T1DM onset, 88 people with latent autoimmune diabetes in adults and 119 people with type 2 diabetes; Results: ZnT8 autoantibody positivity was found in 18% of autoantibody-positive schoolchildren, ZnT8 autoantibodies were found in 56% of people with T1DM, ZnT8 autoantibodies were detected in 10% of people with latent autoimmune diabetes in adults; Conclusion: “ZnT8 autoantibodies are useful markers for prediction of type 1 diabetes in a general population”.

#### Source 13

Reference: Rogowicz-Frontczak A, et al.^29^; Objective: to assess the prevalence ZnT8 autoantibodies, other diabetes-related autoantibodies and clinical manifestation of type 1 diabetes in adults; Type of study: cross-sectional study; Participants: 119 patients with T1DM; Results: 45.4% T1DM < 35 and 34% T1DM ≥ 35 subjects were positive for ZnT8 autoantibodies; Conclusion: “ZnT8 autoantibodies positivity is related to higher title and more frequent occurrence of multiple diabetes-related”.

#### Source 14

Reference: Gomes KF, et al.^30^; Objective: to evaluate the relevance of ZnT8A for T1DM diagnosis; Type of study: case-control study; Participants: 629 patients with T1DM and 651 controls; Results: Znt8A was detected in 68.7% of recent-onset T1DM patients and 48.9% of the entire patient cohort; Conclusion: “ZnT8A detection increases T1DM diagnosis rate even in mixed populations”.

#### Source 15

Reference: Kawasaki E, et al.^31^; Objective: to determine the prevalence and role of autoantibodies to ZnT8A in fulminant form, acute-onset form, and slow-onset form of Japanese patients with T1DM; Type of study: cross-sectional; Participants: 196 new-onset patients with T1DM, 85 fulminant, 81 acute-onset, and 30 slow-onset T1DM type 1; Results: ZnT8A were detected in 58% patients with acute-onset and 20% with slow-onset type 1 diabetes; Conclusion: “ZnT8A are an additional useful marker for acute-onset T1DM”.

#### Source 16

Reference: Garnier L, et al.^32^; Objective: Evaluate the added value of screening anti-ZnT8 antibodies for the diagnosis of T1DM within a large cohort of both children and adults; Type of study: Retrospective 2-year study; Participants: 516 patients (215 children, 301 adults); Results: 110 patients were ZnT8A-positive; Conclusion: “ZnT8A should be included in routine evaluation at diabetes onset and is a valuable biological marker to classify newly-diagnosed diabetics”.

#### Source 17

Reference: Andersen MK, et al.^33^; Objective: to assess the prevalence of ZnT8 autoantibodies in patients with adult-onset diabetes; Type of study: cross-sectional; Participants: 264 T1DM and 294 latent autoimmune diabetes in adults; Results: ZnT8A were significantly more prevalent in latent autoimmune diabetes in adults (34.3%) compared to adult-onset T1DM (18.7%). Conclusion: “ZnT8A were more common and more persistent in patients with latent autoimmune diabetes in adults compared to adult-onset T1DM”.

#### Source 18

Reference: Salonen KM, et al.^34^; Objective: to define the characteristics of humoral autoimmunity against ZnT8 in children and adolescents with newly diagnosed T1DM; Type of study: cross-sectional; Participants: 2,115 subjects <15 years of age; Results: ZnT8 antibodies were detected in 63% of the cases; Conclusion: “Antibodies for ZnT8 is related to age and metabolic status at T1DM diagnosis”.

#### Source 19

Reference: Yang L, et al.^35^; Objective: to evaluate the utility of ZnT8A for diagnosis of autoimmune T1DM in Chinese relative to other autoantibody markers; Type of study: cross-sectional study; Participants: 539 T1DM; Results: ZnT8A were present in 24.1% (130 of 539) of patients with T1DM; Conclusion: “ZnT8A is an independent marker for T1DM in Chinese”.

#### Source 20

Reference: Petruzelkova L, et al.^36^; Objective: to evaluate prevalence of autoantibodies to ZnT8 in Czech children at the onset of T1DM; Type of study: case-control study; Participants: 227 children with newly diagnosed Type 1 diabetes and from 101 control children without diabetes; Results: ZnT8 autoantibodies were detected in 163/227 (72%) of children at T1DM onset and in 1/101 (1%) of the control subjects. Conclusion: “Measurements of ZnT8 autoantibodies are important for Type 1 diabetes diagnosis and should be included in the panel of autoantibodies tested at the onset of T1DM”.

#### Source 21

Reference: Fabris M, et al.^37^; Objective: to investigate ZnT8A as a complement to the current T1DM; Type of study: retrospective multicentre study; Participants: 213 T1DM paediatric patients; Results: ZnT8A showed positive results in 106/213 (49.8 %) T1DM patients; Conclusion: “Study confirms ZnT8A as an important additional and independent diagnostic marker of T1DM”.

#### Source 22

Reference: Araujo DB, et al.^38^; Objective: to investigate the prevalence of ZnT8 autoantibodies in patients with T1DM; Type of study: cross-sectional study; Participants: 72 T1DM patients; Results: The prevalence of ZnT8A was of 24%; Conclusion: “ZnT8 autoantibodies is observed in non-Caucasian patients with T1D, even years after the disease onset”.

#### Source 23

Reference: Niechcial E, et al.^39^; Objective: To assess the prevalence of ZnT8 autoantibodies in children and adults with T1DM onset; Type of study: cross-sectional study; Participants: 367 patients (218 children; 149 adults) at the T1DM onset; Results: ZnT8 autoantibodies (81.1%) in youth; Conclusion: “ZnT8 autoantibodies are associated with more acute diabetes onset”.

### Prevalence of ZnT8 antibodies

The studies reported a variable prevalence of ZnT8 antibodies in individuals with T1DM, ranging from 10.00 % to 90.00 %. The prevalence appeared to be influenced by factors such as age, ethnicity, and disease duration.

### Diagnostic accuracy of ZnT8 antibodies

Some of the studies evaluated the sensitivity, specificity, and positive and negative predictive values of ZnT8 antibodies for T1DM diagnosis. The overall diagnostic performance varied across studies.

Some studies explored the association between ZnT8 antibody positivity and specific T1DM subtypes or disease progression.

This heterogeneity may have limited the ability to draw definitive conclusions about the overall diagnostic accuracy of ZnT8 antibodies.

### Methodological Evaluation and Risk of Bias

The quality of the included studies will be assessed using the QUADAS-2 tool (Table 1).

**Tabela 1.**
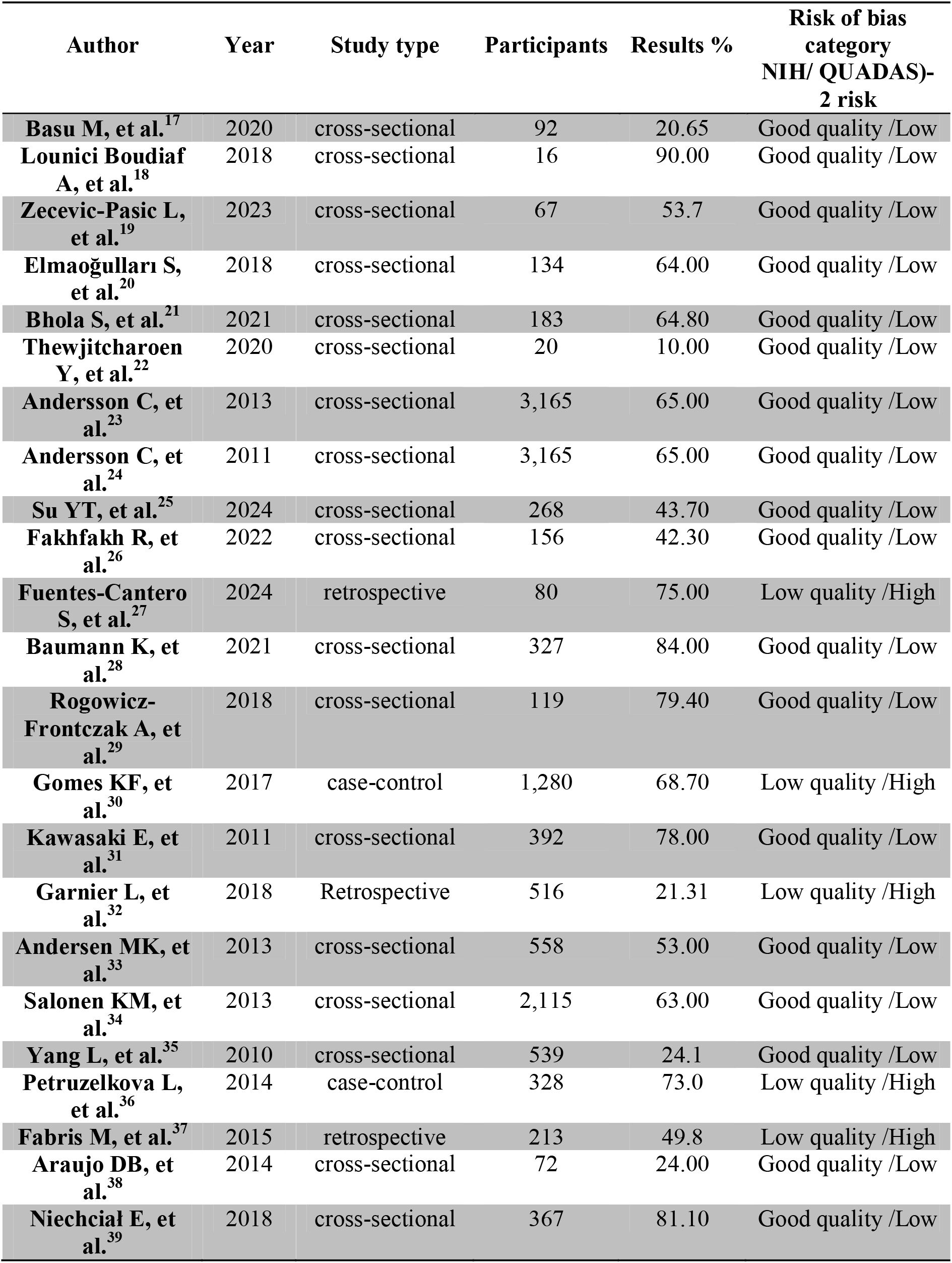
General Characteristics of the Studies Included.

### Meta-Analysis of ZnT8 Autoantibody Positivity and T1DM

This study conducted a comprehensive meta-analysis to evaluate the association between ZnT8 autoantibody positivity and T1DM. We employed a systematic search strategy, encompassing all relevant association studies published between February 2007 and May 2024, identified through a thorough PubMed search.

The analysis yielded a total of 23 original studies investigating the relationship between ZnT8 autoantibodies and T1DM. These studies collectively included a substantial sample size of 14,172 subjects. To quantify the strength of this association, we estimated the relative risk for T1DM using allelic OR.

Reassuringly, the analysis revealed no statistically significant heterogeneity in the genotypic distribution across the included studies. This was confirmed by both the Woolf test (v2 = 43.36, df = 31, P = 0.07) and the Higgins statistic (I^2^ = 28.1%). Additionally, to assess for potential publication bias, we employed the conservative Egger’s regression asymmetry test and found no evidence of bias (P = 0.41).

Given the absence of heterogeneity and publication bias, we opted for a Mantel-Haenszel procedure (fixed effects model) to generate a pooled OR for the association between ZnT8 autoantibody positivity and T1DM. This analysis yielded a statistically significant pooled OR of 1.14 (95% CI: 1.12-1.16 - P = 0.07) (Figure 2).

**Figure 2.**
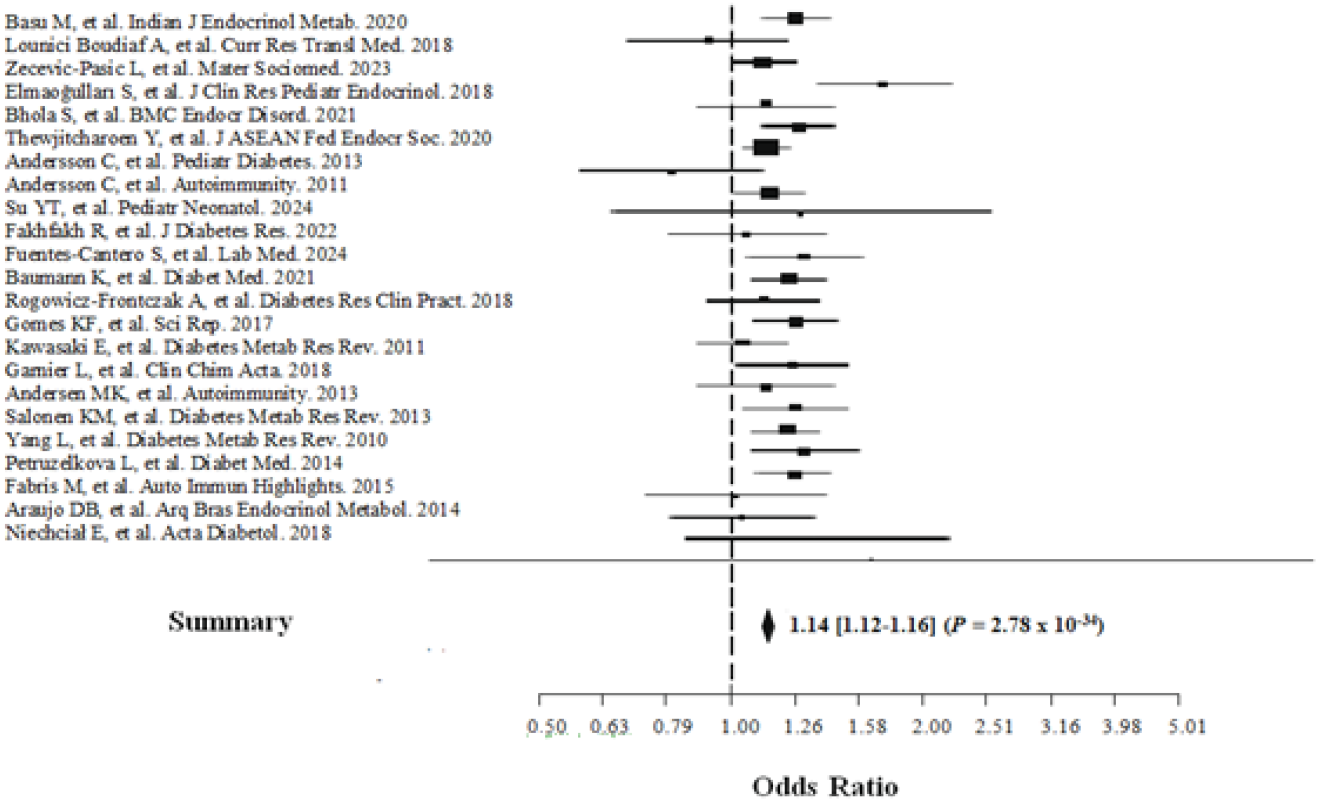
This forest plot depicts the association between the Znt8 gene and T1DM across various studies. Each study is represented by a square. The area of the square reflects the precision of the estimated effect size (OR). A larger square indicates a more precise estimate, stemming from a lower standard error. The horizontal line extending from the square represents the 95% confidence interval (CI) for the effect size. The overall association across studies is summarized by a diamond. The width of the diamond is inversely proportional to its standard error, again conveying the level of precision. The horizontal edges of the diamond represent the lower and upper limits of the pooled 95% CI for the summary OR.

### Limitations

The studies included in this review exhibited heterogeneity in terms of study design, participant characteristics, and assay methods used for ZnT8 antibody detection.

## DISCUSSION

Our systematic review and meta-analysis provides robust evidence supporting a significant association between anti-ZnT8 autoantibody positivity and an increased risk of T1DM development. In this study, we investigate the potential of ZnT8 autoantibody positivity as a diagnostic marker for T1DM, a chronic autoimmune condition characterized by pancreatic β-cell destruction. Given the established link between autoimmunity and increased susceptibility to other chronic immune disorders, a comprehensive evaluation of ZnT8’s diagnostic accuracy in T1DM is warranted. This knowledge may prove instrumental in the development of future prophylactic and therapeutic strategies. To this end, we conducted a meticulous systematic review and meta-analysis, employing a rigorously defined search strategy across pertinent databases. By leveraging well-established Boolean operators and a curated selection of keywords, we aimed to extract the most relevant data on ZnT8’s diagnostic efficacy in T1DM.

Prior investigations exploring the link between ZnT8 autoantibody positivity and T1DM yielded conflicting results.^40^ To address this heterogeneity, we conducted a comprehensive meta-analysis encompassing 22 studies. This analysis revealed a statistically significant elevation in ZnT8 positivity rates within T1DM cohorts compared to control groups, particularly within case-control studies. Notably, despite inherent variations amongst the included studies, analyses demonstrated a lack of undue influence from any single study on the overall meta-analysis OR. Collectively, these findings underscore the robustness and reliability of the conclusions drawn from this meta-analysis, strengthening the evidence for ZnT8 as a potential diagnostic biomarker in T1DM.

This systematic review underscores the value of routine islet autoantibody screening, particularly anti-ZnT8 antibodies, in identifying individuals at risk of developing T1DM. While a previous meta-analysis advocated for a similar approach,^41^ the present study further strengthens the body of evidence supporting the role of ZnT8 antibodies in T1DM diagnosis. This, in turn, enhances our understanding of the contribution of autoimmunity to T1DM development in susceptible populations.

Early identification of T1DM cases enables prompt intervention to mitigate the risk of diabetic ketoacidosis and other T1DM-related complications,^20,42^ as corroborated by a systematic review highlighting the increased risk of autoimmune comorbidities among T1DM patients.^17^ Additionally, these strategies can also improve metabolic control.^41^

This systematic review and meta-analysis are not without limitations. Firstly, the observed heterogeneity among the included studies, potentially arising from variations in sample size, warrants cautious interpretation of the findings. Additionally, despite the substantial sample size, pooled estimates adjusted for covariates could not be extracted. To our knowledge, this represents the first systematic review and meta-analysis investigating the association between ZnT8 autoantibodies and T1DM in a diabetic population.

Despite these limitations, our comprehensive systematic review and meta-analysis provide compelling evidence supporting the robust association between anti-ZnT8 autoantibody positivity and an increased risk of T1DM development. The findings from this meta-analysis underscore the value of anti-ZnT8 autoantibodies as a diagnostic biomarker for T1DM, offering a promising tool for early identification and intervention strategies.

## CONCLUSION

Our systematic review and meta-analysis provides robust evidence supporting a significant association between anti-ZnT8 autoantibody positivity and an increased risk of T1DM development. Additionally, studies investigating the cost-effectiveness of routine islet autoantibody screening, including anti-ZnT8 antibodies, are needed to inform clinical practice guidelines.

## Data Availability

All data produced in the present work are contained in the manuscript

## Conflicts of interest

The authors declare no conflicts of interest.

## Notes

### Competing Interest Statement

The authors have declared no competing interest.

### Funding Statement

This study did not receive any funding

### Author Declarations

The study used (or will use) ONLY openly available human data that were originally located at: Pubmed

